# Clinician belief revision after a background electroencephalography biomarker report in a nurse-led first seizure clinic: the PRISTINE study

**DOI:** 10.1101/2024.12.13.24318654

**Authors:** Phil Tittensor, Alan M Batterham, Jacqui-Dawn Rowe, Daniel Russell, Danielle Steward, Elizabeth Schnabel, Kay Meiklejohn, Milaana Mainstone, Hamish Myers, Benjamin B Howes, Yolanda R Hill, Lydia E Staniaszek, Rosie Charles, Shaun Wellburn, John R Terry, Wessel Woldman

**Affiliations:** The Royal Wolverhampton NHS Trust, Wolverhampton, UK; University of Wolverhampton, Wolverhampton, UK; Professor Emeritus, School of Health and Life Sciences, Teesside University, Middlesbrough, UK; The Royal Wolverhampton NHS Trust, Wolverhampton, UK; Neuronostics, Bristol, UK; Neuronostics, Bristol, UK & University Hospitals Bristol and Weston NHS Foundation Trust, Bristol, UK; Neuronostics, Bristol, UK; University Hospital Southampton NHS Foundation Trust, Southampton, UK; Neuronostics, Bristol, UK. Shaun Wellburn: South Tees Hospitals NHS Foundation Trust, Middlesbrough, UK; Neuronostics, Bristol, UK; University of Plymouth, Plymouth, UK; Neuronostics, Bristol, UK; University of Birmingham, Birmingham, UK

**Keywords:** epilepsy diagnosis, observer variation, diagnostic decision support, clinical decision-making, diagnostic uncertainty, probability elicitation

## Abstract

**Objective:** To measure how clinicians in a first seizure clinic revise their recorded probability of a further epileptic seizure on receiving the report of BioEP, an EEG biomarker computed from background activity, and to explore, post hoc, how the report bands relate to the recorded 12-month working diagnosis.

**Methods:** In a prospective single-site belief-updating study, 86 adults with a suspected first seizure formed the analysis set. A consultant nurse for the epilepsies rated the probability of a further epileptic seizure on seven-category and visual analogue scales after clinical review and routine EEG (step 1), and again after the BioEP report (step 2). An independent, experienced clinical nurse specialist for the epilepsies rated the same probabilities from documentation. All care was decided at step 1; step 2 entries were for research only. Five contributory EEGs were outside the BioEP indication; the 81 non-contributory EEGs (the primary analysis population) comprised 48 directional and 33 uncertain baseline assessments. The 12-month endpoint was the consultant nurse’s recorded working diagnosis, not independently adjudicated.

**Results:** The consultant nurse changed category in 32/81 patients (40%, 95% CI 29 to 51); revision direction and mean magnitude followed the ordered BioEP report bands. In a post hoc exploratory analysis of 33 patients with an uncertain baseline assessment, 17/18 unsupportive BioEP reports were concordant with a non-epilepsy 12-month working diagnosis. Three supportive reports provided no evidence of a rule-in signal. As an exploratory whole-cohort pathway comparison against the same working diagnosis, routine EEG followed by BioEP classified as positive 18/24 patients with an epilepsy working diagnosis against 4/24 for routine EEG alone, at specificity 63% and 98% respectively.

**Significance:** BioEP report viewing was associated with graded revision of clinicians’ recorded beliefs. PRISTINE was not designed to establish diagnostic accuracy, calibration, management effect or patient benefit.

**Plain language summary:** After a possible first seizure, EEG often reveals nothing. BioEP is a computer analysis of that normal-looking EEG. It reports whether it looks more like, or less like, EEG from people with epilepsy. A consultant nurse for the epilepsies recorded how likely a further seizure seemed, before and after the BioEP report, in 81 adults whose EEG was inconclusive. Their estimate changed in 32 out of 81 patients. When they were uncertain in their diagnosis at the first visit, an unsupportive BioEP result was almost always followed by no recorded epilepsy working diagnosis at 12 months. This requires confirmation.

**Key points:** • The consultant nurse for the epilepsies changed the rated probability category in 32/81 patients after seeing the BioEP report.

• All care was decided before the report was seen; belief revision is the prospective finding.

• Revision direction and mean magnitude followed the ordered BioEP report bands for both raters.

• In 33 uncertain baseline assessments, 17/18 unsupportive BioEP reports were concordant with a non-epilepsy working diagnosis.

## Introduction

Epilepsy is a clinical diagnosis. A multi-factorial process including the history and any witness account, alongside investigations (of which the routine EEG is the most widely used) informs the decision [1,2]. The routine EEG is specific but insensitive: pooled sensitivity after a first unprovoked seizure is 17% at specificity 95% [3], and more than 70% of routine EEGs show no epileptiform abnormality in adults [2]. Since a routine EEG without epileptiform abnormality does not exclude epilepsy [1], most patients leave a first seizure clinic with the EEG having contributed nothing to their diagnosis. As a result, diagnostic delay beyond a year is common and misdiagnosis in adults is estimated at 20 to 30% [4–6]. At the end of the clinical visit, the clinician therefore has not a binary result, but rather a degree of suspicion (effectively a working diagnosis) which is revised as further evidence accumulates. The nurse-led clinic where this study took place has reported outcomes for its first 200 patients: 92% received a working diagnosis at the first visit and 74% of EEGs were reported as normal [7].

Experts differ in reading the EEG and interpreting the clinical history. For example, 35 US neurologists reviewing the same 200 routine EEG segments marked between 6 and 212 epileptiform discharges each [8]. Furthermore, when 154 physicians with an interest in epilepsy judged a defined set of case studies in two stages, agreement on whether the person had epilepsy rose from a kappa of 0.56 (1 being perfect agreement) on the clinical description to 0.70 once the results of investigations were added to the clinical description [9].

Collectively, these studies suggest that clinicians form and revise diagnostic beliefs in the absence of a common standard, such that the working diagnosis recorded depends on the individual clinician who assessed the patient.

To provide additional context for clinical decision making, BioEP is an EEG biomarker for epilepsy computed from background activity rather than from epileptiform discharges. Eight predefined features, two spectral, four describing the functional network derived from the recording and two derived from a mathematical model of that network, are combined by a pre-trained statistical model into a single score, reported in five ordered bands from Very Unsupportive to Very Supportive of a diagnosis of epilepsy [10–13]. The computation is deterministic. In a retrospective multicentre case-control study using two-tier cross-validation, BioEP applied to 281 clinically non-contributory recordings from eight NHS sites gave balanced accuracy of 68% (sensitivity 61%, specificity 75%) [10]. A further retrospective analysis of 196 non-contributory EEGs from six NHS sites has since examined its use in prioritising follow-up [14]. BioEP is UKCA-marked as a Class I medical device. Its context of use is diagnostic decision support for adults with suspected epilepsy and a non-contributory routine EEG.

Crucially, retrospective classifier performance cannot describe what happens when clinical judgement is augmented by a decision support tool such as BioEP. The interaction can, though, be measured directly. Where a clinician records an estimate of the probability of a further epileptic seizure before and after seeing new evidence, the change in that estimate expresses the evidential weight placed on it, and the implied change in odds can be recovered from the ratings themselves [15]. The estimated probability of a further seizure can therefore be used to quantify the clinician’s diagnostic belief and its revision, rather than serving as a validated prognostic prediction.

We report the findings of the PRISTINE study, which applied this design in a nurse-led first seizure clinic. Its registered primary outcome was the change in the consultant nurse for the epilepsies (CN) rating, recorded after clinical review and routine EEG and again after the BioEP report. An experienced epilepsy specialist nurse (ESN) acted as an independent second rater. They recorded the same judgements, so that the effect could be observed in more than one clinician. All care was decided at the first assessment, and the BioEP report did not inform decisions. Belief revision is therefore the only prospectively measured effect. We additionally report exploratory analyses involving the 12-month outcome. Reporting follows GREENBEAN guidelines, with DECIDE-AI guidelines informing the reporting of clinician-tool interaction and human factors [16,17].

## Methods

### Design, setting and governance

PRISTINE was a prospective, single-site diagnostic belief-updating study conducted in shadow mode in a live clinic: the BioEP report was generated and viewed but did not affect care. It was sponsored and funded by Neuronostics Ltd, registered before recruitment (ClinicalTrials.gov NCT05764252) and approved by the West Midlands, Solihull Research Ethics Committee (IRAS 321340; REC reference 23/WM/0083). The setting was a CN-led first seizure clinic at The Royal Wolverhampton NHS Trust: a one-stop clinic where most patients have a routine EEG at the visit, reported at the time by a consultant clinical neurophysiologist [7] (see Supplementary Material).

### Registered design, implemented conduct and follow-up endpoint

Protocol version 1.0 (1 March 2023) and the ClinicalTrials.gov registration anticipated that the BioEP score might inform clinical decisions, and registered longitudinal outcomes including additional investigations and services, decisions over repeated presentations, and BioEP analysis of any follow-up EEG. As ultimately implemented, every clinical decision was made and recorded at step 1, before the report was seen, and the step 2 fields for medication, investigation and referral were completed for research purposes only and did not alter care. A crosswalk of registered outcomes against those collected, analysed and reported is given in Supplementary Table S2. The registered longitudinal outcomes presupposed that BioEP could influence care. Since it could not, those outcomes are not reported here.

The 12-month endpoint was the CN’s working diagnosis recorded at 12 months after the index visit, classified as epilepsy or not epilepsy by applying the ILAE practical definition [18]. The working diagnosis was made and maintained by the same CN who made the baseline assessment, through the clinic’s usual reviews at around three, six to nine, and twelve months after the index visit [7]. The 12-month endpoint was not independently adjudicated, and the second rater’s assessments (the rater is described under Belief measurement) informed neither clinical care nor the 12-month endpoint. Because the same clinician provided both the baseline rating and the working diagnosis used as the endpoint, agreement between the two is described as concordance and cannot be interpreted as an independent measure of diagnostic accuracy. In addition, where the clinician held a directional assessment at baseline, any subsequent working diagnosis was not independent of that initial assessment [19,20].

Furthermore, whilst fixing management at step 1 meant the BioEP report could not alter the initial care decision, the clinician retained knowledge of the report during follow-up.

Therefore, later influence on follow-up intensity or diagnostic revision cannot be excluded. One adjudication rule was applied: a participant assessed as non-epileptic at the index presentation re-attended months later with different symptoms. A sleep-deprived EEG requested for those symptoms was contributory and epilepsy was diagnosed. The participant was withdrawn at close-out due to a new diagnosis emerging from a separate presentation and not the index event.

### Participants

Eligible patients were adults aged 18 or over presenting with a first suspected seizure, able to give informed consent, and referred for a routine EEG. The protocol excluded patients unable to tolerate an EEG, patients with known hepatic or renal encephalopathy, and patients in whom the history gave a clear diagnosis of a physical condition other than epilepsy. BioEP was used in accordance with its instructions for use, which exclude recordings from patients with structural abnormality, including skull defect. All eligible patients attending the clinic between 23 August 2023 and 12 August 2024 who were offered a routine EEG were approached, and none declined. Demographic, lifestyle, medication, comorbidity, risk-factor and index-event data were recorded at the visit. The planned sample of 88 was the maximum feasible in one year, giving a 95% confidence interval width of about 0.20 around the primary proportion.

### Index test

The index test was BioEP V1.0.0 (Neuronostics, Bristol, UK). The routine EEG (minimum 19 channels, 10-20 placement) was exported in EDF format, pseudonymised, and uploaded by the neurophysiology team. The participant’s sex and anti-seizure medication status, both required by BioEP, are provided. The platform identifies artefact-free segments of eyes-closed background activity, computes the eight predefined features described above and combines them through the pre-trained model to give the score expressed as one of five ordered bands: Very Unsupportive, Unsupportive, Neutral, Supportive and Very Supportive. The feature set, model and band boundaries were fixed before recruitment and remained unaltered during the study. The report displays the band, the score with its confidence interval, the proportion of epilepsy and non-epilepsy cases in the development database that fell in that band, and a fixed caution against use as a standalone test (Supplementary Figure S1). The routine EEG was classified from the neurophysiologist’s report as contributory (epileptiform abnormality), normal non-contributory, or abnormal non-contributory (non-specific abnormality only); BioEP is indicated only in cases where the EEG is non-contributory.

### Belief measurement and procedure

At step 1, after clinical review of the routine EEG report and any other investigation available at the visit, the CN rated the probability of a further epileptic seizure on a seven-category scale adapted from the IPCC guidance on communicating uncertainty [21], with about as likely as not (33% to less than 66%) at its centre. Additionally, a 100-mm visual analogue scale was used for the same purpose. At step 2, the CN opened the BioEP report and repeated both ratings. Throughout, ’band’ refers to the BioEP report and ’category’ to the raters’ seven-point scale. An independent ESN acted as second rater, completing both scales at steps 1 and 2 from the documented history and investigation reports, without patient contact and with no access to the CN’s ratings or clinic correspondence. Both clinicians received instructions for use and training on the report and scale reporting during site initiation visit.

### Analysis populations

The exploratory pathway comparison uses all 86 patients, the five contributory EEGs representing routine EEG alone; every other analysis, including the primary outcome, uses the 81 patients with a non-contributory EEG, for whom BioEP is indicated. Patients were classified according to their step 1 rating. The “clinician-directional” group comprised those rated at step 1 in any category other than “about as likely as not”, whereas the “clinician-uncertain” group comprised those rated “about as likely as not”. The “clinician-uncertain” group was recorded prospectively at step 1; the restriction of the patient-level follow-up analysis to this group was post hoc.

## Statistical analysis

Statistical analyses used Stata (Release 19) and Python (3.11) with SciPy. The primary outcome was the proportion of patients in whom the CN’s rating moved by at least one category between steps 1 and 2, with a 95% Clopper-Pearson exact confidence interval as pre-specified [22]. Wilson score intervals are used for the exploratory analyses, which hold nearer nominal coverage at small sample sizes [23]. The same proportion was derived for the second rater and the visual analogue scale. Within-rater odds multipliers implied by the ratings, defined as post-report odds divided by pre-report odds, were calculated from category midpoints and, separately, from the visual analogue values. These were summarised as medians with interquartile ranges for revisions towards and away from epilepsy [15].

Revisions were classified by direction relative to the BioEP report, a revision after a Neutral report being counted separately because that band indicates no direction, and their size was described by band using Spearman’s rank correlation. The protocol’s exploratory question, whether revision was more likely when the report agreed with the prior [24], is addressed descriptively. Revision in the uncertain and directional groups is described but not formally compared, because the direction of any difference depends on the scale used.

Post data collection, three exploratory analyses involving the 12-month working diagnosis were specified. At the pathway level, in the full cohort, routine EEG alone was compared with routine EEG followed by BioEP on the same recording. A patient counted as positive if the EEG was contributory or, where non-contributory, if the BioEP band was not Unsupportive or Very Unsupportive. This dichotomy follows the product’s labelling, since Very Unsupportive and Unsupportive weigh against epilepsy and the other bands do not, and is applied as a binary cut point; the binary analysis was not pre-specified, and the boundary was compared with data-derived optima. Because the combined strategy nests the contributory EEGs, each difference between strategies reduces to a single proportion with a Wilson interval; Newcombe’s paired method was a robustness check (Supplementary Material) [25].

At the patient level, in the clinician-uncertain group, the proportion of unsupportive results with a non-epilepsy working diagnosis was calculated with its two-sided 95% Wilson interval and tested against the group base rate (observed prevalence) with a one-sided exact binomial test at α = 0.025, matching the two-sided 95% interval, unadjusted for multiplicity.

At the rater level, the second rater’s discrimination of the working diagnosis before and after the BioEP report was compared by the area under the receiver operating characteristic curve and the Brier score, with paired bootstrap confidence intervals. The registered medication outcome was analysed on the step 1 recommendation, tabulated by band with Spearman’s correlation and Fisher’s exact test between the band ends, with and without patients taking a medication at presentation. Missing second-rater ratings were excluded without imputation.

## Usability and patient and public involvement

Report co-design and summative usability evaluation under IEC 62366-1:2015, conducted as remote pre-deployment simulation, are described in the Supplementary Material [17,26,27]. Members of the South West chapter of Epilepsy Action reviewed the rationale, protocol and participant materials before submission for approval.

## Results Participants

Of the 91 patients consented, five were withdrawn or removed: two could not tolerate an EEG of analysable length, one consent was not valid, one was withdrawn at close-out under the adjudication rule, and one was removed for a skull defect, an exclusion under the instructions for use not identified at screening (Figure 1). The analysis set was 86 patients, forming four nested groups: five contributory EEGs outside the indication, and 81 non-contributory EEGs comprising 48 directional and 33 uncertain baseline assessments. Across the four groups, the recorded 12-month working diagnosis was epilepsy in 24 of 86 (28%), 20 of 81 (25%), 10 of 48 (21%) and 10 of 33 (30%). All 86 recordings returned a BioEP result; second-rater ratings were missing for three patients at one or both steps. Baseline characteristics are in Table 1.

**Figure 1.**
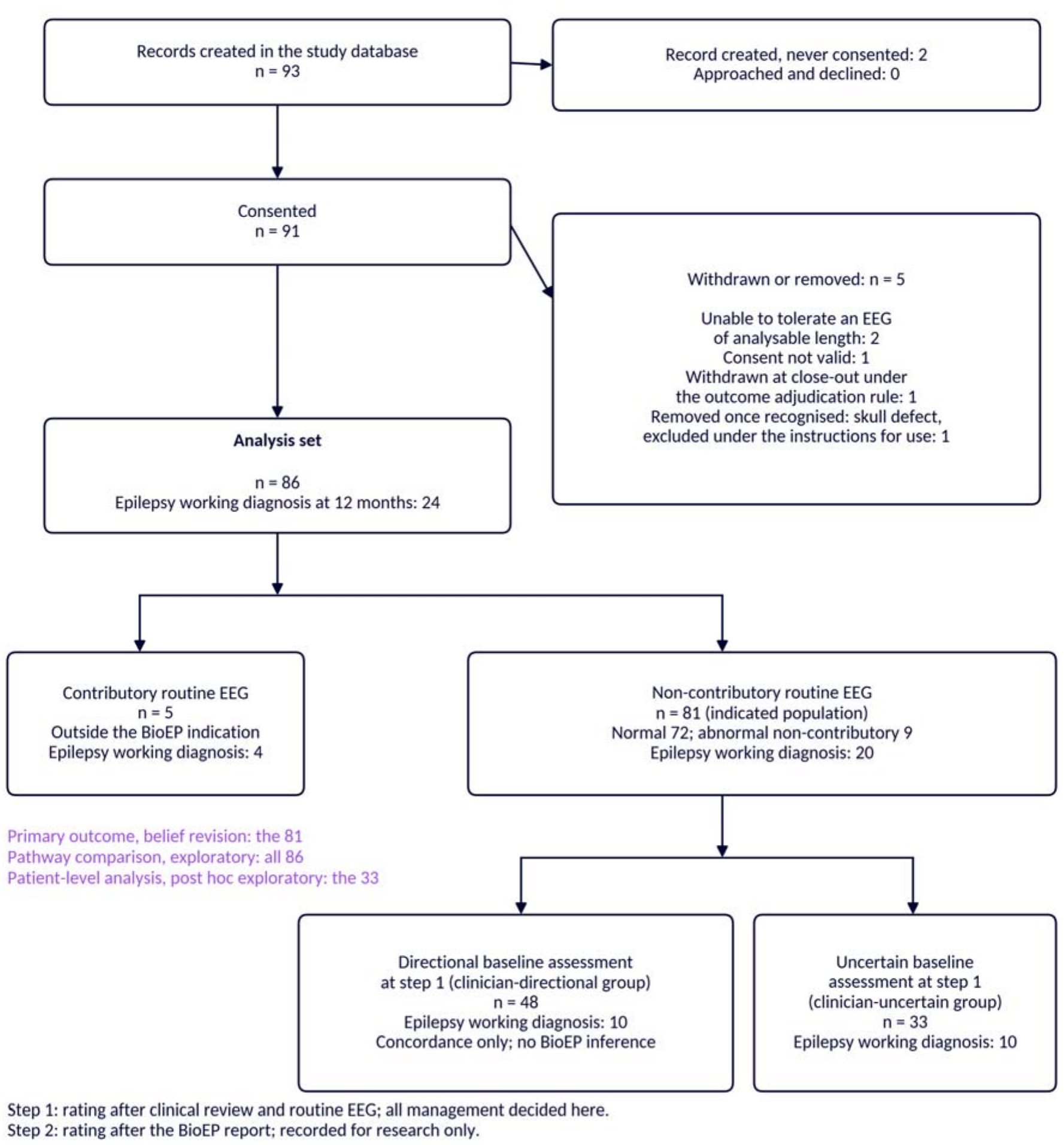
Participant flow. Step 1 is the consultant nurse’s rating after clinical review and routine EEG, at which point all management was decided; step 2 is the rating after the BioEP report, recorded for research only. The outcome throughout is the consultant nurse’s recorded 12-month working diagnosis.

**Table 1.** Baseline characteristics of the analysis set, by analysis population. Values are n (%) unless stated. Sex is sex at birth; gender was also recorded and differed for two participants. Smoking and alcohol missing for four, recreational drug use for one, risk-factor form not completed for one. Comorbidities were free text; "no comorbidity recorded" counts entries of none, nil or blank. Risk factors and triggers were multiple-choice, so rows are not mutually exclusive. Pregabalin prescribed for pain or anxiety was not counted as an anti-seizure medication.

| **Characteristic** | **All (86)** | **Indicated (81)** | **Clinician-uncertain (33)** | **Directional (48)** |
|---|---|---|---|---|
| Age, years, median (IQR) | 36 (24 to 60) | 36 (25 to 60) | 48 (29 to 61) | 34 (24 to 60) |
| Male sex | 44 (51) | 41 (51) | 22 (67) | 19 (40) |
| Current smoker | 12 (14) | 12 (15) | 7 (21) | 5 (10) |
| Ex-smoker | 20 (23) | 20 (25) | 4 (12) | 16 (33) |
| Alcohol over 10 units per week | 21 (24) | 19 (23) | 8 (24) | 11 (23) |
| Recreational drug use | 9 (10) | 8 (10) | 3 (9) | 5 (10) |
| No comorbidity recorded | 28 (33) | 25 (31) | 12 (36) | 13 (27) |
| Anti-seizure medication at presentation | 10 (12) | 10 (12) | 2 (6) | 8 (17) |
| Family history of epilepsy | 20 (23) | 19 (23) | 7 (21) | 12 (25) |
| Head injury | 9 (10) | 9 (11) | 4 (12) | 5 (10) |
| No risk factor recorded | 47 (55) | 43 (53) | 17 (52) | 26 (54) |
| Index event trigger unknown | 47 (55) | 46 (57) | 20 (61) | 26 (54) |

## Belief revision

The CN’s category moved in 32 of 81 patients (40%; 95% CI 29 to 51), towards less likely in 19 and towards more likely in 13, in every case by a single category (Table 2). On the visual analogue scale the value changed in 56 of 81 (69%). The independent second rater’s category moved in 49 of 78 rated patients (63%; 51 to 74), by one category in 43 and by two or three in six. Revisions followed the BioEP report: of the CN’s 32 category moves, 24 were in the direction the report indicated, one against it, and seven occurred where the BioEP report read “Neutral”. On the visual analogue scale, 42 of the 44 changes made when the BioEP report was not “Neutral” were in its direction.

**Table 2.** Belief revision between step 1 and step 2, by rater. The consultant nurse saw the patient; the independent epilepsy specialist nurse rated from the documented history and investigation reports. Odds multipliers are post-report odds divided by pre-report odds, from category midpoints. Second-rater ratings were missing for three patients at one or both steps. Category midpoints: 0.5, 5.5, 22, 50, 78, 94.5 and 99.5%. Visual analogue multipliers and mean change by band are in Supplementary Table S3.

|  | **Consultant nurse (n=81)** | **Second rater (n=78)** |
|---|---|---|
| Category moved by at least one, n (%; 95% CI) | 32 (40; 29 to 51) | 49 (63; 51 to 74) |
| Moved towards epilepsy / away from epilepsy | 13 / 19 | 19 / 30 |
| Category moves: in the report's direction / against it / where the report read Neutral / all moves | 24 / 1 / 7 / 32 | 33 / 3 / 13 / 49 |
| Visual analogue value changed, n (%) | 56 (69) | 54 (69) |
| Odds multiplier, revisions away from epilepsy: median (IQR), n | 0.28 (0.21 to 0.28), 19 | 0.28 (0.21 to 0.28), 30 |
| Odds multiplier, revisions towards epilepsy: median (IQR), n | 4.9 (3.6 to 11.6), 13 | 4.9 (4.9 to 8.2), 19 |

Mean change in the visual analogue rating followed the BioEP band ordering, from -12 mm after Very Unsupportive to +18 mm after Very Supportive for the CN (Spearman’s rho 0.82, p<0.001) and from -17 to +48 mm for the second rater (rho 0.67) (Figure 2). The implied within-rater odds multipliers were modest and roughly symmetric: on the category scale, the CN’s median multiplier was 0.28 for revisions away from epilepsy and 4.9 towards it (Table 2; per-band values in Supplementary Table S3).

**Figure 2.**
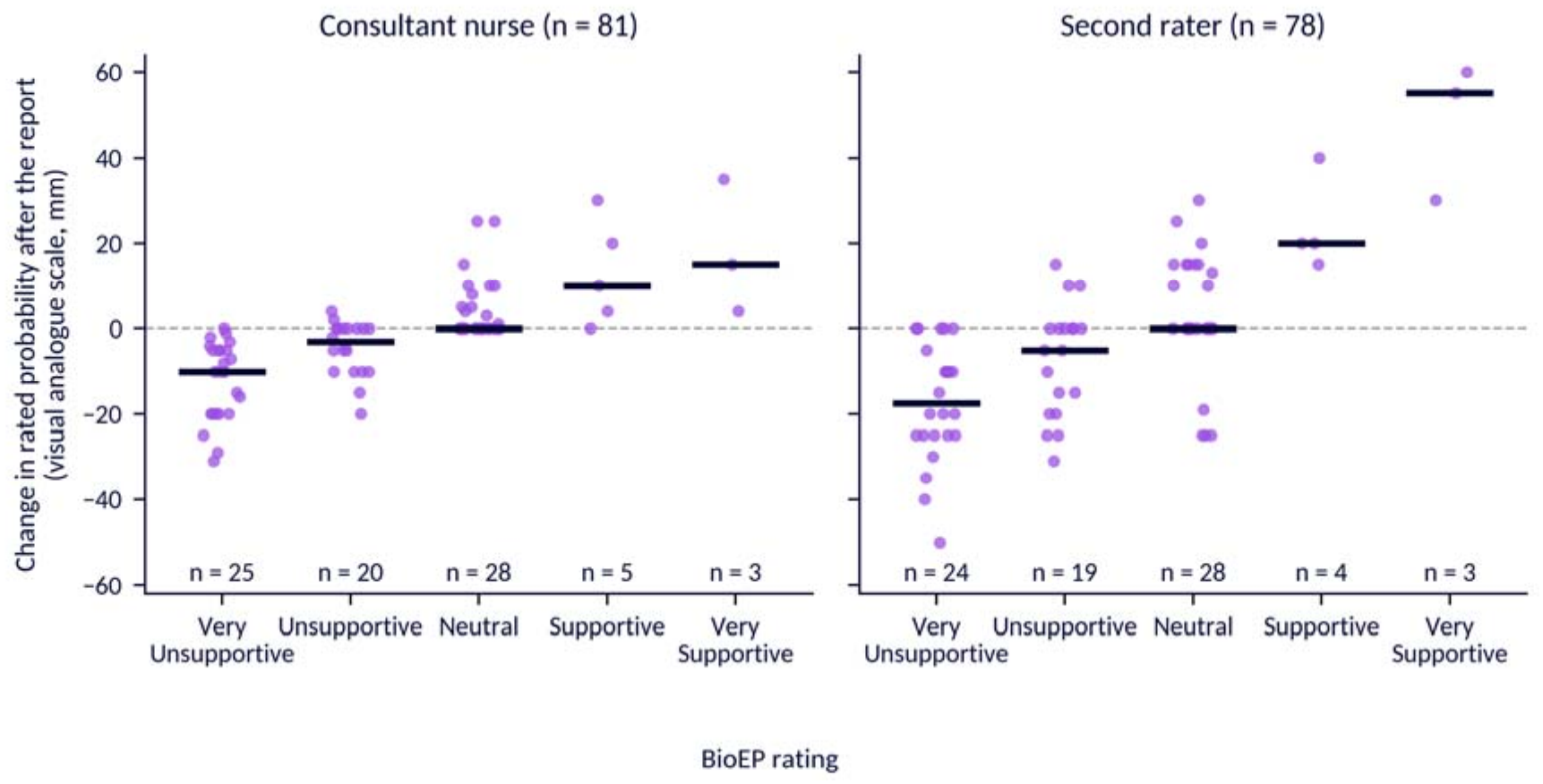
Change in the rated probability of a further epileptic seizure on the visual analogue scale between step 1 and step 2, by BioEP band, for the consultant nurse (left) and the independent second rater (right), in the 81 patients within the indication. Each point is one patient; horizontal bars are medians.

Revision was not confined to reports that agreed with the prior. Where the BioEP report pointed against a directional step 1 rating, the CN’s visual analogue value moved in 12 of 14 patients, and where the BioEP report agreed, the visual analogue value moved in 14 of 21 patients.

## Concordance in the clinician-directional group

Of the 48 patients with a directional baseline assessment, the step 1 direction matched the recorded 12-month working diagnosis in 45: 13 were rated towards epilepsy, of whom 10 carried an epilepsy working diagnosis. None of the 35 who were rated away from epilepsy had a working diagnosis of epilepsy. This is concordance rather than accuracy, because the baseline assessment shaped both the management and the working diagnosis; no interval, accuracy measure or BioEP inference is drawn from it.

## Analysis in the 33 clinician-uncertain patients

In post hoc exploratory analysis of the 33 patients with an uncertain baseline assessment, 17 of 18 unsupportive reports were concordant with no epilepsy in the CN’s recorded 12-month working diagnosis (Table 3). The endpoint remained non-independent, since the same clinician saw the report and provided follow-up care. The observation therefore suggests a potential rule-out role for an unsupportive report in this subgroup but requires confirmation in an independent cohort. The three supportive reports provide no evidence of a rule-in signal. After the report, the CN revised the visual analogue value towards less likely in 16 of the 33, of whom one carried an epilepsy working diagnosis at 12 months.

**Table 3.** Post hoc exploratory analysis: BioEP band against the. consultant nurse’s recorded 12-month working diagnosis in the 33 patients with an uncertain baseline assessment at step 1. *Unsupportive or Very Unsupportive, 17 of 18 with a non-epilepsy working diagnosis (94%; 95% CI 74 to 99), against 12.5 expected at the 30% base rate (one-sided exact p=0.013, unadjusted for multiplicity or post hoc selection, as an exploratory analysis). Supportive or Very Supportive: one of three with an epilepsy working diagnosis (p=0.66)*.

| **BioEP band** | **Patients** | **Epilepsy working diagnosis** | **No epilepsy working diagnosis** |
|---|---|---|---|
| Very Unsupportive | 11 | 0 | 11 |
| Unsupportive | 7 | 1 | 6 |
| Neutral | 12 | 8 | 4 |
| Supportive | 1 | 0 | 1 |
| Very Supportive | 2 | 1 | 1 |
| Total | 33 | 10 | 23 |

## Pathway comparison in all 86 patients

Against the same non-independent working diagnosis, routine EEG alone classified positive four of the 24 patients with an epilepsy working diagnosis (17%; CI 7 to 36) at specificity 98% (CI 91 to 100), while routine EEG followed by BioEP at the deployed band boundary classified positive 18 of 24 (75%; CI 55 to 88) at specificity 63% (CI 50 to 74) (Supplementary Table S1). The combined strategy classified positive an additional 14 of the 24 who went on to receive an epilepsy working diagnosis (58 percentage points; CI 39 to 76), at a specificity cost of 36 points (CI 25 to 48), an incremental yield of 14 of 86 (16 points; CI 10 to 26). The band boundary coincided with the Youden and Liu optima in this dataset. Of the 45 patients negative on both, six had an epilepsy working diagnosis: one in the clinician-uncertain group and five in the directional group.

## Rater level analysis

In a post hoc exploratory analysis, the second rater’s discrimination of the working diagnosis was higher after the report (n=78; area under the curve 0.93 against 0.86 before; difference +0.06, 95% CI -0.02 to +0.14; Brier score 0.108 against 0.125).

## Registered medication outcome

At step 1, before the report was seen, the CN recommended an anti-seizure medication in 19 of the 81 indicated patients, and the proportion did not track the BioEP report band (7/25, 5/20, 4/28, 2/5 and 1/3 across the five bands; Spearman’s rho -0.05, p=0.64; unsupportive end 12 of 45 against supportive end 3 of 8, Fisher’s exact p=0.67). Excluding the 10 patients already taking an anti-seizure medication at presentation, who accounted for 9 of the 19 recommendations, a medication was recommended in 10 of 71 (5/23, 2/17, 2/25, 1/4 and 0/2). These are recommendations recorded at the visit, not confirmed treatment uptake. BioEP was unseen at step 1, so this is a design-fidelity check: the acted-on recommendation preceded the report and was therefore unrelated to it.

## Discussion

PRISTINE progresses beyond the retrospective evidence of classifier performance [10] to address the clinically consequential question: what does a biomarker report add to the judgement of a clinician who already has access to the patient’s history? After routine clinical review and routine EEG, the CN changed the recorded probability category in 32 of 81 patients on seeing the BioEP report, and revision followed the ordered BioEP bands on both rating scales.

The second rater, working from documentation without a patient encounter, showed the same gradient. Two raters holding different information, each moving in proportion to the same ordered scale, cannot be attributed to expectation alone and supports the report being weighed as evidence. The odds multipliers implied by the ratings were modest, symmetric and graded by band, and revision occurred regardless of whether the report agreed with the prior; the report was neither ignored nor deferred to. The exploratory rater-level analysis indicates that the revisions moved in the direction the outcomes supported.

The findings point to where that evidential weight may be most useful. In the 33 patients whose baseline assessment was uncertain, 17 of 18 unsupportive reports were concordant with a recorded non-epilepsy working diagnosis at 12 months, a post hoc association that suggests a rule-out role but requires confirmation in an independent cohort. Its interest lies in the setting: most routine EEGs after a suspected first seizure contain no epileptiform abnormality [2], and these are the patients for whom diagnosis takes longest [6]. A biomarker capable of providing evidence against epilepsy when routine EEG is non-contributory and clinical assessment remains uncertain could therefore address a point in the diagnostic pathway at which existing clinical investigations provide limited information. An exploratory whole-cohort comparison at the deployed band boundary shows the trade-off a confirmatory study will need to quantify: the additional patients classified as positive who went on to receive an epilepsy working diagnosis came at a cost in specificity (incremental yield increase: 16 percentage points), so a Neutral result should prompt further evaluation rather than treatment (Supplementary Table S1). A double negative did not exclude epilepsy, but five of the six such cases had a clinician already leaning towards epilepsy, consistent with the report contributing most where the clinician was uncertain.

Most prediction models for recurrence after a first seizure take EEG abnormality as an input [28,29], and automated review tools identify epileptiform activity on the contributory EEG [30]. BioEP addresses the non-contributory EEG and is complementary to them.

## Limitations

The study is single site, with one CN of significant experience and one second rater, so generalisation is untested, and a more sceptical clinician might revise less. Routine EEG was reported by one department, through four consultant clinical neurophysiologists, without central reading. The 12-month endpoint is the CN’s recorded working diagnosis rather than independent adjudication. The same CN saw the report and provided follow-up, so cognitive carry-over into follow-up intensity or diagnostic revision cannot be excluded. Further, restriction to uncertain baseline assessments reduces, but does not remove, incorporation of a directional judgement. The study therefore does not establish diagnostic accuracy, whether the size of each revision was appropriate, or a safe basis for de-escalating care. Because no management decision could be influenced, the study could assess neither safety nor clinical benefit. The primary outcome is a self-reported belief with no external referent: re-rating immediately after the report may itself have prompted revision. The uncertain-group restriction was post hoc, with five tests reported without adjustment for multiplicity due to the exploratory nature of the analyses. The lower limit of the uncertain-group interval is compatible with about one in four unsupportive results having epilepsy. The Neutral band was allocated to the positive side because its label does not weigh against epilepsy; treating it as neither would also be consistent with that label. Because the binary pathway analysis was not pre-specified and its boundary coincided with the in-sample optimum, its estimates may be optimistic; no threshold is proposed for clinical use.

## Confirmation

Confirmation of the rule-out findings requires a prospective study in which a rule-out endpoint is pre-specified, the report is available to inform management, and outcomes are determined independently of the clinician whose belief is measured. Two design points carry forward: showing that epilepsy is less common after an unsupportive result than at the base rate is a modest ask, whereas showing that a rule-out threshold clears a level high enough to change management needs a far larger study, with the operating point locked before any prospective data are seen.

## Conclusion

In a nurse-led first seizure clinic, viewing the BioEP report was followed by revision of the recorded probability of a further epileptic seizure in 40% of patients, in the direction and to a degree that followed the ordered BioEP report bands, while all care remained fixed. A post hoc exploratory analysis of patients with an uncertain baseline assessment found that 17 of 18 unsupportive reports were concordant with a recorded non-epilepsy working diagnosis, generating a rule-out hypothesis that should be tested independently in future work.

## Author contributions (CRediT)

Conceptualisation: PT, AMB, KM, MM, SW, JRT, WW. Methodology: PT, AMB, HM, BBH, YRH, LES, RC, SW, JRT, WW. Investigation: PT, JDR, DR, DS, ES, HM. Data curation: KM, MM. Formal analysis: AMB, SW, JRT. Visualisation: AMB, JRT, WW. Writing, original draft: PT, AMB, JRT, WW. Writing, review and editing: all authors. Supervision: PT, JRT, WW. Project administration: KM, MM. Funding acquisition: JRT, WW.

## Supporting information

DECIDE-AI checklist

GREENBEAN checklist

## Data Availability

The EEG recordings and meta-data are not publicly available due to restrictions by privacy laws. Post-processed data supporting the findings of this study are available upon reasonable request from the corresponding author (WW), and additional meta-data may be made available upon reasonable request from the study sponsor (MM).

## Acknowledgements

We thank the neurophysiology and epilepsy nursing teams of The Royal Wolverhampton NHS Trust, the members of the South West chapter of Epilepsy Action, led by Simon Privett, who reviewed the study materials, and Professor Greg Atkinson for independent statistical peer review of the protocol.

## Ethics Statement

The study was approved by the Health Research Authority and Health and Care Research Wales (IRAS 321340) and by the West Midlands, Solihull Research Ethics Committee (reference 23/WM/0083), and was conducted in accordance with the Declaration of Helsinki and Good Clinical Practice. All participants included in the analysis gave written informed consent; one record was removed because consent had been given by a relative and was not valid.

## Use of artificial intelligence-assisted technologies

The large language model Claude (Anthropic, San Francisco, USA) was used to assist with drafting, editing and restructuring the manuscript, to check reported values for consistency against the study dataset, and to write the code with which the exploratory analysis was performed. The study statistician (AMB) performed the primary and pathway analyses independently of the model and reviewed the exploratory analyses. The authors reviewed and edited all content and take full responsibility for it. Generative AI was not used in the design or conduct of the study or in data collection. BioEP itself does not use generative AI.

## Data availability

The EEG recordings and clinical data are not publicly available because of privacy and ethical restrictions. Post-processed, de-identified data supporting the findings are available on reasonable request from the corresponding author, and the analysis scripts from the study statistician or JRT (depending on the specific analysis). The BioEP feature set and modelling approach are published [10–13]; the deployed implementation is proprietary. Its technical documentation is available to regulators.

