## Supplementary Material for "Clinician belief revision after a background electroencephalography biomarker report in a nurse-led first seizure clinic: the PRISTINE study"

Running title: PRISTINE belief-updating study

**Authors**

Phil Tittensor*, Alan M Batterham*, Jacqui-Dawn Rowe, Daniel Russell, Danielle Steward, Elizabeth Schnabel, Kay Meiklejohn, Milaana Mainstone, Hamish Myers, Benjamin B Howes, Yolanda R Hill, Lydia E Staniaszek, Rosie Charles, Shaun Wellburn, John R Terry†, Wessel Woldman†

*Joint first authors. †Joint senior authors.

Affiliations. Phil Tittensor: The Royal Wolverhampton NHS Trust, Wolverhampton, UK; University of Wolverhampton, Wolverhampton, UK. Alan M Batterham: Professor Emeritus, School of Health and Life Sciences, Teesside University, Middlesbrough, UK. Jacqui-Dawn Rowe, Daniel Russell, Danielle Steward, Elizabeth Schnabel: The Royal Wolverhampton NHS Trust, Wolverhampton, UK. Hamish Myers: Neuronostics, Bristol, UK. Benjamin B Howes: Neuronostics, Bristol, UK. Yolanda R Hill: Neuronostics, Bristol, UK. Lydia E Staniaszek: Neuronostics, Bristol, UK & University Hospitals Bristol and Weston NHS Foundation Trust, Bristol, UK. Kay Meiklejohn: Neuronostics, Bristol, UK; University Hospital Southampton NHS Foundation Trust, Southampton, UK. Milaana Mainstone: Neuronostics, Bristol, UK. Rosie Charles: Neuronostics, Bristol, UK. Shaun Wellburn: South Tees Hospitals NHS Foundation Trust, Middlesbrough, UK. John R Terry: Neuronostics, Bristol, UK; University of Plymouth, Plymouth, UK. Wessel Woldman: Neuronostics, Bristol, UK; University of Birmingham, Birmingham, UK.

**Supplementary material**

**EEG specifics**

EEG was recorded with a Cadwell system running ARC software, with a sampling rate of 500 Hz, in referential montage (reference electrode: Cz). Activation methods included intermittent photic stimulation and hyperventilation for 3 minutes. EEGs were read by experienced consultant clinical neurophysiologists (ANS and AHCS registered).

**Pathway comparison**

This was an exploratory analysis. Sensitivity, specificity, predictive values and yield were calculated for routine EEG alone and for routine EEG followed by BioEP, against the CN's recorded 12-month working diagnosis. The combined strategy counted a patient as positive if the routine EEG was contributory or, where non-contributory, if the BioEP band was other than Unsupportive or Very Unsupportive; BioEP therefore contributed only for the 81 non-contributory recordings. The dichotomy follows the product's labelling but was not pre-specified as an analysis, and it coincides with the Youden and Liu optima in this dataset, so estimates for the combined strategy and for the differences between strategies may be optimistic. The combined strategy is a nested superset of the contributory EEGs: no patient could be positive on routine EEG and negative on the combined strategy, which was confirmed in the dataset. Because of this nesting, the paired difference in sensitivity reduces to a single proportion, the number of patients with an epilepsy working diagnosis who were negative on routine EEG and positive on the combined strategy as a fraction of all patients with an epilepsy working diagnosis, and carries a Wilson score interval. The same logic applies to the loss in specificity and to the incremental yield, the latter as a fraction of all 86 patients. As a robustness check that does not rely on the nesting, each difference was also estimated with Newcombe's Method 10 for paired proportions. Wilson intervals were used throughout, except the positive predictive value of routine EEG alone, where the denominator of five calls for an exact interval. Predictive values are reported descriptively rather than as a formal difference. Intervals are two-sided at 95% and conditional on the cut point. Analyses were performed in Stata Release 19.

**Figure S1.** Exemplar BioEP report (not based on real patient data).

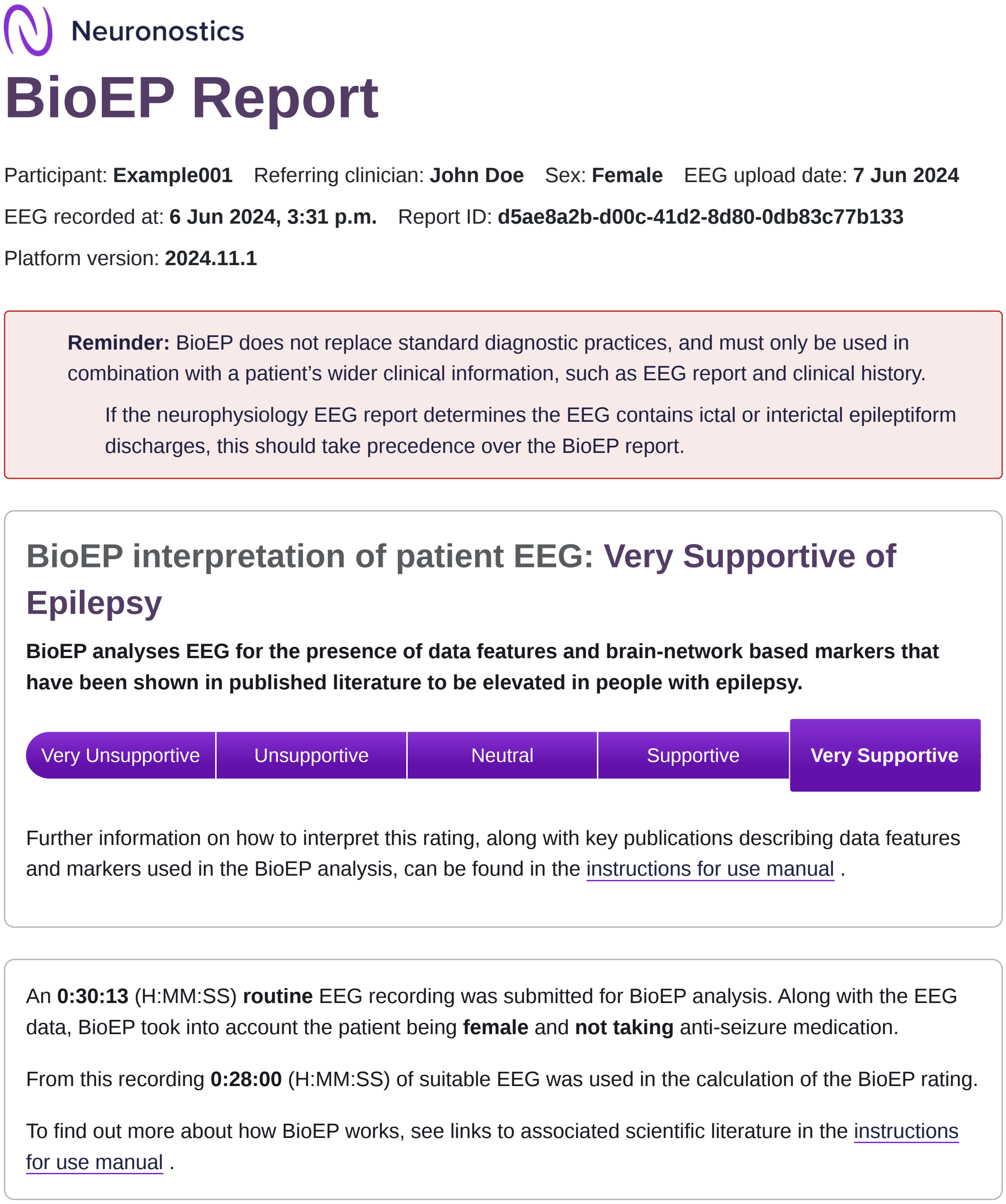

Neuronostics

### BioEP Report

Participant: **Example001** Referring clinician: **John Doe** Sex: **Female** EEG upload date: **7 Jun 2024**

EEG recorded at: **6 Jun 2024, 3:31 p.m.** Report ID: **d5ae8a2b-d00c-41d2-8d80-0db83c77b133**

Platform version: **2024.11.1**

**Reminder:** BioEP does not replace standard diagnostic practices, and must only be used in combination with a patient's wider clinical information, such as EEG report and clinical history.

If the neurophysiology EEG report determines the EEG contains ictal or interictal epileptiform discharges, this should take precedence over the BioEP report.

#### BioEP interpretation of patient EEG: Very Supportive of Epilepsy

**BioEP analyses EEG for the presence of data features and brain-network based markers that have been shown in published literature to be elevated in people with epilepsy.**

Very Unsupportive | Unsupportive | Neutral | Supportive | **Very Supportive**

Further information on how to interpret this rating, along with key publications describing data features and markers used in the BioEP analysis, can be found in the instructions for use manual .

An **0:30:13** (H:MM:SS) **routine** EEG recording was submitted for BioEP analysis. Along with the EEG data, BioEP took into account the patient being **female** and **not taking** anti-seizure medication.

From this recording **0:28:00** (H:MM:SS) of suitable EEG was used in the calculation of the BioEP rating.

To find out more about how BioEP works, see links to associated scientific literature in the instructions for use manual .

**Table S1.** Exploratory pathway comparison against the consultant nurse's recorded 12-month working diagnosis, all 86 patients: routine EEG alone against routine EEG followed by BioEP at the deployed band boundary.

| Measure | Routine EEG alone | Routine EEG then BioEP | Difference, percentage points (95% CI) |
|---|---|---|---|
| Sensitivity, % (n/N) | 16.7 (4/24) | 75.0 (18/24) | +58.3 (38.8 to 75.5) |
| Specificity, % (n/N) | 98.4 (61/62) | 62.9 (39/62) | -35.5 (-47.9 to -24.7) |
| Yield, % of all 86 (n/N) | 4.7 (4/86) | 20.9 (18/86) | +16.3 (10.0 to 25.5) |
| Positive predictive value, % (n/N; 95% CI) | 80.0 (4/5; 28.4 to 99.5) | 43.9 (18/41; 29.9 to 59.0) | not compared |
| Negative predictive value, % (n/N; 95% CI) | 75.3 (61/81; 64.9 to 83.4) | 86.7 (39/45; 73.8 to 93.7) | not compared |

A patient counts as positive if the routine EEG was contributory or, where non-contributory, if the BioEP band was Neutral, Supportive or Very Supportive. Because the combined strategy nests the contributory EEGs, each difference reduces to a single proportion (14/24, 22/62 and 14/86) and carries a Wilson interval; Newcombe's method for paired proportions gave the same point estimates with slightly wider intervals, mainly at the lower limit: sensitivity gain 32.5 to 73.7, specificity reduction 22.7 to 48.0, and incremental yield 8.4 to 25.4. Intervals are Wilson except the positive predictive value of routine EEG alone, which is exact given the denominator of five. Predictive values are reported per strategy rather than as a difference because the five contributory EEGs limit any comparison. The prevalence of an epilepsy working diagnosis was 38.9% (24.8 to 55.1) among the 36 patients negative on routine EEG and positive on BioEP, against 27.9% (19.5 to 38.2) overall. Of the 45 patients negative on both, six (13.3%) had an epilepsy working diagnosis: one in the clinician-uncertain group and five in the directional group. The cut point was not pre-specified and coincides with the Youden and Liu optima, so estimates may be optimistic; all intervals are conditional on the cut point.

**Table S2. Registered and protocol-anticipated outcomes against outcomes collected, analysed and reported**

| Outcome (source) | Collected | Analysed | Reported |
|---|---|---|---|
| Clinician's perception of seizure probability, two-step, seven-category and visual analogue scales, with implied odds multipliers (registry primary; protocol 3.2) | Yes | Yes | Yes (primary outcome) |
| Decisions to recommend commencing or deferring anti-seizure medication (registry; protocol 4.3) | Yes (step 1 decision; step 2 research-only field) | Yes | Yes (registered medication outcome) |
| Number of additional investigations or services (registry; protocol 4.3) | No | No | No |
| Clinicians' decisions over repeated presentations (registry; protocol 3.3) | No | No | No |
| BioEP analysis of follow-up EEGs in the 12-month period, with the score presented to the clinician (registry; protocol 3.3, 4.3) | No | No | No |
| Cognitive bias in belief updating: revision when the report agreed or disagreed with the prior (protocol 3.3) | Yes | Yes (descriptive) | Yes (results, belief revision) |
| Definitive diagnosis and medication decision within 12 months (protocol 3.3) | Yes (working diagnosis) | Yes (exploratory) | Yes, as the CN's recorded 12-month working diagnosis |

Protocol section 7.1 and the ClinicalTrials.gov registration anticipated that decisions might be affected by the BioEP score. As implemented, all care was decided at step 1 and step 2 entries were research-only.

**Table S3. Belief revision detail by rater**

| | **Consultant nurse (n=81)** | **Second rater (n=78)** |
|---|---|---|
| Moved by one category / by two or more | 32 / 0 | 43 / 6 |
| Visual analogue moved up / down | 21 / 35 | 21 / 33 |
| Mean visual analogue change by band, Very Unsupportive to Very Supportive, mm | -12, -4, +4, +13, +18 | -17, -7, +3, +24, +48 |
| Spearman's rho, band against visual analogue change | 0.82 ($p<0.001$) | 0.67 |
| Odds multiplier, visual analogue scale, revisions away from epilepsy: median (IQR), n | 0.52 (0.37 to 0.67), 35 | 0.33 (0.33 to 0.44), 33 |
| Odds multiplier, visual analogue scale, revisions towards epilepsy: median (IQR), n | 3.4 (1.9 to 5.2), 21 | 3.0 (3.0 to 8.1), 21 |
| Step 1 rating about as likely as not, n | 33 of 81 | 22 of 81 (17 of the 33) |

Visual analogue values of 0 or 100 (six ratings) were set to 0.5 and 99.5 for the odds calculation. Eight second-rater category-value disagreements were reconciled to the numeric value before analysis.

**Co-design and usability evaluation of the BioEP report**

Rationale. A decision support tool is a sociotechnical intervention: its effect depends on how the user reads and weighs its output. Usability engineering for medical devices is governed by IEC 62366-1:2015 [1,2]. The hazards of particular relevance to a probabilistic output are misinterpretation of the result and reliance on the result without the clinical context; both were carried through formative and summative evaluation, in line with guidance on co-designing clinical practice tools [3] and with the DECIDE-AI requirement to report human factors evaluation and user familiarisation [4]. The work was remote, pre-deployment simulation. It did not assess live workflow, learning curves, clinical effectiveness or patient outcomes.

Formative evaluation. Eight clinicians from clinical sites and academic institutions took part in remote, quasi-structured sessions with early versions of the report. The cohort included consultant neurologists (n=4), a consultant clinical neurophysiologist, a clinical physiologist, a consultant nurse for the epilepsies, and a consultant neuropsychiatrist. Participants were identified using convenience sampling and purposive recruitment through direct outreach (using NHS frameworks and websites), and the sessions took place between 13th of January 2023 and 10th of March 2023. These resulted in four changes: the layout was simplified to foreground the band and its interpretation; the category labels were rephrased as support for a diagnosis rather than its likelihood; the graphical presentation of the score was adjusted; and the caution against reliance on the score alone was made more prominent.

Summative evaluation. Fifteen intended users from clinical sites and academic institutions interpreted the report for pseudonymised cases in structured remote simulation sessions built around the two hazard scenarios. The cohort included consultant neurologists (n=7), consultant clinical neurophysiologists (n=2), specialist nurses (n=4), and consultant neuropsychiatrists (n=2). Participants were identified using convenience sampling and purposive recruitment through direct outreach, and the sessions took place between 11th of October 2023 and 15th of November 2023. All 15 completed the primary tasks and no use error occurred that could lead to an unacceptable risk of harm.
