## Supplementary material for "Clinician belief revision after a background electroencephalography biomarker report in a nurse-led first seizure clinic: the PRISTINE study": GREENBEAN checklist

Checklist from Ewen et al. 2025; applicability notes are taken from the checklist (Table 4) and from Table 5 (variations by study design).

Study design factors (GREENBEAN Table 5): spontaneous (resting) recording; longitudinal (12-month follow-up); prospective; no intervention; single site; binarised outcome; multimodal.

Validation phase: a prospective single-site study with Phase 3 design features but does not meet all Phase 3 requirements.

| Section & Topic | No | Item | Applicability Notes | Reported on page |
|---|---|---|---|---|
| **TITLE OR ABSTRACT** | | | | |
| | **1** | Identify "biomarker," "EEG." Specify Context of Use and phase of validation | | p1; p2 |
| **INTRODUCTION** | | | | |
| | **2** | State unmet clinical need to be addressed and context of Use. Explain the limitations of current "standard of care" methods of assessment | | pp4-5 |
| | **3** | Justification for EEG features studied | | p5 |
| **METHODS** | | | | |
| *Study Design* | **4** | Description of the study design, including any longitudinal data collection and presence or absence of intervention | | pp6-7 |
| *Participants* | **5** | Inclusion/exclusion criteria, ascertainment/recruitment procedures (including prospective/retrospective design), stratification procedures (if applicable), and relationship among them | | pp7-8 |
| | **6** | Describe how the data were used (e.g., for training and validation/test) in the analysis, considering sample size requirements | Phases 2-4 only | p8 |
| | **7** | Prevalence of diagnosis/outcome in training and test samples or distribution of clinical independent variable | | p11 |
| | **8** | Justification of control group | When different samples are recruited separately | N/A (single prospectively recruited cohort) |
| *Test Methods* | **9** | Index test, in sufficient detail to allow replication | | p8 |
| | **9a** | Participant behavioral management | | p8 |
| | **9b** | Hardware and software, including electrode montage, electrode reference, and recording settings | | p8, Supplementary Material p2 |
| | **9c** | Recording environment | | p6 |

| Section & Topic | No | Item | Applicability Notes | Reported on page |
|---|---|---|---|---|
| | **9d** | Technologist qualifications/training/certification | | N/A (standard qualifications) |
| | **9e** | Psychophysical equipment, tasks, order, and instructions for participants | Not relevant for spontaneous recordings (Table 5) | N/A (spontaneous recording) |
| | **9f** | Calibration of stimulus-delivery devices | | N/A (no stimulus delivery) |
| | **9g** | Data analysis pipeline in sufficient detail to allow replication, including preprocessing of EEG data, data selection/rejection, approaches/criteria/parameters | | p8 |
| | **9h** | Quality control procedures, including staff training and inter-site calibration | Irrelevant to single-site studies (Table 5) | N/A (single site) |
| | **10** | Disclosure of all clinical information or results available to personnel involved in data collection and analysis (both index and reference tests), specifying if they were blinded to certain data | | pp7-9 |
| | **11** | Pre-specification of EEG dependent variables, analysis pipeline and thresholds, and disclosure of all investigator-set parameters or steps explored/revised after data collection began | Required for test samples of Phases 3-4 | p8; p10; p17; Supplementary Material p2 |
| | **12** | Reference standard, its rationale, and procedures for determining alternative diagnoses/outcomes, in sufficient detail to allow replication | | p7 |
| | **13** | Criteria for any exclusion of any participants following (attempted) data collection | | pp7-8; p11 |
| *Statistical Analysis* | **14** | All other risk, disease and demographic variables analyzed alongside EEG dependent variables | | p8; p11; Table 1, p13; p17 |
| | **15** | For multivariable prediction models, details of any variable selection procedures, data-reduction strategies, and other model-building issues | For studies that include multivariable models | p5, p8; model building in references 10-13 |
| | **16** | How missing data and outliers were handled and practical reasons for any missing values | | p11; p12; Table 1, p13; Supplementary Material p7 |
| | **17** | Methods for estimating or comparing measures of diagnostic accuracy, disease progression, etc. | | pp9-11; Supplementary Material p2 |
| | **18** | Details of any correction for multiple comparisons | For Phases 1-2 or the training samples of Phase 3 | p11; Table 3, p16; p20 |
| **RESULTS** | | | | |
| *Participants* | **19** | Flow of participants, using a diagram; including details on participants who did not tolerate testing, did not have interpretable data or withdrew from the study (including explanation why they did so) | | Figure 1, p12; p11 |
| | **20** | All factors relevant to clinical outcome and that can influence the EEG result, including baseline demographic and clinical characteristics of participants, reporting differences between samples (disease/control or training/test) | | p11; Table 1, p13 |

| Section & Topic | No | Item | Applicability Notes | Reported on page |
|---|---|---|---|---|
| | **21** | Time interval and any clinical interventions between index test and reference standard | | pp6-7; pp17-18 |
| | **22** | Change in any clinical variables over the course of a longitudinal study | Irrelevant to concurrent studies (Table 5) | p7 |
| *Test Results* | **23a** | Estimates of accuracy and their precision (such as confidence intervals) | | Table 3, pp16-17; Supplementary Table S1, p4 |
| | **23b** | Results demonstrating the added value of EEG biomarker beyond other data sources known to the clinician | Phases 3-4 | pp13-17 (Table 2, pp14-15) |
| | **24** | For training sample, ROC curve by classification threshold | For some Phase 2 and all Phase 3 (training sample) studies that employ binarized outcomes | N/A (no training sample) |
| | **25** | Graphical visualization of relevant variability and/or consistency in the EEG output (e.g., ERP grand average with confidence intervals, split-half plots or butterfly plots) | | N/A (computational analysis of non-contributory EEG) |
| | **26** | For samples where the prevalence of relevant independent variables (e.g., diagnosis prevalence) or covariates (e.g., age) are dissimilar to the typical clinical population of interest on whom the biomarker will be employed, estimates of population PPV/NPV | | Supplementary Table S1, p4 |
| **DISCUSSION** | | | | |
| | **27** | Study limitations, including sources of potential bias/confounding, statistical uncertainty, and generalizability | | pp19-20 |
| | **28** | Practical implementation factors, including cost of implementation, training and certification of staff, quality control, stigma, need for special testing environments, etc. | | p9 |
| | **29** | Feasibility of all tasks for the clinical population | | p11; Supplementary Material pp8-9 |
| | **30** | Anticipated or demonstrated impact on patient outcomes | Requirement for Phase 4A | pp18-19 |
| | **31** | Demonstration that biomarkers contribute information beyond other sources of (demographic, clinical/disease) information that will be known by clinician | | pp13-18 |
| **OTHER CONSIDERATIONS** | | | | |
| | **32** | Provide details about the availability of analysis code, software and data | | pp22-23 |
| | **33** | Sources of funding and other support; role of funders | | p21 |
| | **34** | Use of generative AI in the production of the manuscript | | p22 |
